# Three Cerebellar Imaging Subtypes in Schizophrenia with Distinct Spatiotemporal Trajectories and Biological Characteristics

**DOI:** 10.64898/2026.10.03.26364634

**Authors:** Zhaoyun Liu, Zhenyu Huang, Xinjia Lin, Jingyu Zhou, Hao Hu, Qian Guo, Yingying Tang, Tianhong Zhang, Jijun Wang, Weihua Yue, Yuyanan Zhang, Xin Yu, Long-Biao Cui, Xiao Chang, Huan Huang, Cheng Luo, Dezhong Yao, Ryota Hashimoto, Junya Matsumoto, Kiyotaka Nemoto, Tilo Kircher, Florian Thomas-Odenthal, Paula Usemann, Lea Teutenberg, Benjamin Straube, Igor Nenadić, Frederike Stein, Nina Alexander, Andreas Jansen, Hamidreza Jamalabadi, Nooshin Javaheripour, Jannik Lepper, Udo Dannlowski, Dominik Grotegerd, Susanne Meinert, Kira Flinkenflügel, Rebekka Lencer, Michael Ziller, Edith Pomarol-Clotet, Raymond Salvador, Andriana Karuk, Gianfranco Spalletta, Fabrizio Piras, Daniela Vecchio, Nerisa Banaj, Ali Saffet Gonul, Aslı Ceren Hinc, Kang Sim, Qian Hui Chew, Yann Quidé, Melissa J. Green, Young-Chul chung, Woo-Sung Kim, Soyolsaikhan Odkhuu, Felice Iasevoli, Giuseppe Pontillo, Andrea de Bartolomeis, Sirio Cocozza, Annarita Barone, Arturo Brunetti, Mariateresa Ciccarelli, Mario Tranfa, Tamsyn E Van Rheenen, Susan L Rossell, Matthew Hughes, Will Woods, Sean Carruthers, Philip J. Sumner, Elysha Ringin, Georgia Caruana, Scott R. Sponheim, Jessica A. Turner, Theo G.M. van Erp, Lena Palaniyappan, Paul M. Thompson, Jianfeng Feng, Yuchao Jiang

**Author notes:** Corresponding author: Yuchao Jiang, Correspondence address: School of Biomedical Engineering, Shenzhen University Medical School, Shenzhen University, Shenzhen, PR China.

## Abstract

The potential role of the cerebellum in the pathophysiology of schizophrenia has received relatively insufficient attention. Prior studies characterized cerebellar abnormalities mainly at the group level, without fully considering the heterogeneity of the disease. Here, we applied a machine learning approach (Subtype and Stage Inference, SuStaIn) to cross-sectional 3D volumetric MRIs of the cerebellum, including a total of 3,550 individuals with schizophrenia (1423 females; mean age: 32.3 ± 12.0 years) and 5,771 healthy controls (HC) (2,861 females; mean age: 32.5 ± 12.9 years), from 34 sites worldwide. SuStaIn identified three distinct spatiotemporal trajectories in cerebellar gray matter volume reduction, respectively originating in lobule X (subtype 1), lobule III (subtype 2) and lobule VIIb (subtype 3), with subtypes 1 and 3 corresponding to the posterior lobe and subtype 2 to the anterior lobe. These trajectories were cross-sectionally inferred in 1,588 individuals with schizophrenia and replicated in another independent samples of 1,334 and 628 patients, respectively. Multimodal analyses using neuroimaging, transcriptomic and behavioral data revealed subtype-specific biological characteristics in brain morphological patterns, cerebellar-cortical connectivity, gene expression and clinical symptoms. Specifically, subtype-related genes were enriched in metabolism-related processes and immunity-related processes, respectively, in subtype 1 and subtype 2; subtype 3 showed more severe brain abnormalities and worse cognitive symptoms. Treatment data from 381 patients, with up to 12 months of follow-up, revealed poorer response to antipsychotic medications in subtype 2 and subtype 3, but better response to transcranial magnetic stimulation in subtype 1, which also had worse emotion-related symptoms. Together, our findings offer a comprehensive characterization of the heterogeneity of cerebellar pathophysiological progresses in schizophrenia, which may help in developing future clinical stratification and targeted intervention strategies.

## Introduction

Studies of the cerebellum are systematically under-represented in neuroscience ^[1]^, and there is a lack of knowledge regarding its possible role in schizophrenia, a highly heterogeneous psychiatric disorder involving large-scale abnormalities in the brain network ^[2, 3]^. Prior neuroimaging studies in schizophrenia have mainly focused on the cerebral cortex and subcortical structures such as the hippocampus and basal ganglia, but have paid limited attention to cerebellar pathophysiology in schizophrenia ^[1]^. Recent magnetic resonance imaging (MRI) studies showed reduced gray matter volume (GMV) in cerebellum in individuals with schizophrenia ^[4]^. Research has also demonstrated brain dysfunctions in schizophrenia, implicating abnormalities in cerebello-thalamo-cortical circuitry ^[5, 6]^. At the microscopic level, a neonatal phencyclidine model of schizophrenia found disrupted synaptic connections of cerebellum, together with reduction in Purkinje cells and granule cells, which suggest abnormal integrity of cerebellar neural circuits ^[7]^. Furthermore, cerebellar dysfunction has been related to worsening of negative symptoms ^[8, 9]^ and persistent cognitive impairment in schizophrenia ^[10, 11]^. Taken together, these findings suggest that the cerebellum may be integrally involved in the pathophysiological processes of schizophrenia.

However, prior studies have typically focused on group level differences in cerebellar dysfunction, without fully considering the heterogeneity of the disease. There is considerable individual variability in brain structure and functional connectivity in schizophrenia ^[12, 13]^. Recently, several international Enhancing Neuro Imaging Genetics through Meta-Analysis (ENIGMA) studies have reported increased variability in brain structure among patients, relative to healthy controls (HC), supporting the presence of considerable neuroanatomical heterogeneity in schizophrenia ^[14, 15]^. Clinical symptoms show high individual variability and may exhibit dynamic fluctuations at different disease stages, even for the same patient ^[16]^. Therefore, pathological progresses within the cerebellum may not be explained by a single pathophysiological process, but rather by multiple distinct pathophysiological trajectories, including contributions from abnormalities in the cerebellum.

Traditionally, the cerebellum has been regarded primarily to as coordinating and influencing low-order functions of motor coordination. However, recent studies have argued that the cerebellum is also deeply involved in high-order functions, such as cognitive and emotional processing ^[17–19]^. Importantly, the differences from low-order to high-order functions are not uniformly distributed across the structure but are organised along an anterior-to-posterior gradient axis within the cerebellum ^[20, 21]^. Anterior lobules (I–V) are preferentially connected to primary sensorimotor cortices; the central lobules (VI, Crus I) are associated with higher-order cognitive processes including executive function and working memory; and the posterior lobules (Crus II, Lobule X) are functionally coupled to limbic and default-mode networks and implicated in affective regulation and social processing ^[8, 19]^. This topographic organisation offers a principled framework to interpret the heterogeneity of cerebellar pathology in schizophrenia, and generates testable predictions about the clinical correlates of pathological trajectories originating at different points along this gradient.

Novel machine learning methods define distinct subgroups based on symptom characteristics and neuroimaging indicators ^[22–26]^. Prior studies showed that brain structural abnormalities in schizophrenia patients were not static but showed progressive trajectories, such as widespread thinning of cerebral cortex ^[27]^ and decreased subcortical volume ^[28]^. However, previous studies revealed subtypes in exclusively phenotype ^[29, 30]^ or disease stage ^[31]^, without considering both. This limitation has direct clinical consequences. For instance, if distinct cerebellar pathophysiological trajectories confer differential responsiveness to antipsychotic medications versus brain stimulation interventions (as the present data indicate) then failure to characterise this heterogeneity at the individual level limits the development of stratified treatment strategies, representing a substantive gap in the emerging precision psychiatry agenda for schizophrenia. A modelling framework capable of simultaneously inferring subtype identity and disease stage from cross-sectional neuroimaging data is therefore particularly well-suited to addressing this question. The subtype and stage inference (SuStaIn) algorithm ^[32]^ is precisely such a data-driven disease progression model, which can infer subtype-specific progression patterns using cross-sectional data alone. Therefore, using the SuStaIn algorithm to characterize the cerebellar pathophysiological trajectories may further help understand the heterogeneity in schizophrenia, as well as providing potentially valuable biomarkers for disease classification.

In this study, we analyzed the heterogeneity of the cerebellum based on a large cohort of 1,588 individuals with schizophrenia and examined several specific questions. First, using the SuStaIn algorithm on cross-sectional data, we identified different trajectories of cerebellar GMV reduction in schizophrenia. Secondly, we assessed the patterns of connectivity with the cerebral cortex associated with different cerebellar subtypes and used resting-state functional magnetic resonance imaging (rs-fMRI) to reveal subtype-specific functional connectivity (FC) patterns. Subsequently, we further utilized spatial transcriptomics data to reveal subtype-relevant gene expression patterns and biological pathways. Finally, we investigated the relationship between each subtype and clinical symptoms and examined the differences in efficacy between the subtypes in antipsychotic medication (APM) or transcranial magnetic stimulation (TMS) treatment. Importantly, based on a 12-month follow-up cohort, we investigated whether these SuStaIn subtypes had predictive value for improvement in clinical symptoms. The overall framework of this study is detailed in **Extended Figure 1**. In summary, the classification results found in this study further explain the heterogeneity of the cerebellar pathophysiological progresses in schizophrenia, which may help in developing future clinical stratification and targeted therapeutic strategies.

## Results

### Three distinct spatiotemporal trajectories of cerebellar volume reduction

We applied the SuStaIn algorithm to cross-sectional MRI data, including a total of 1,588 individuals with schizophrenia (638 females; mean age: 31.9 ± 12.0 years) and 2,341 HC (1,041 females; mean age: 33.3 ± 13.6 years) (**Figure 1a**) (**Supplementary Table 1-2**). Mean z-scores of the schizophrenia group exhibited a moderate degree in GMV reduction for nine cerebellar subregions (range from 0.083 to 0.192) (**Figure 1b**). SuStaIn identified three cross-sectionally inferred progression sequences of the nine subregions (i.e., three “trajectories”) (**Figure 1c**). It characterized three distinct spatiotemporal patterns in GMV reduction for the cerebellum (**Figure 1d**). Specifically, trajectory 1 showed GMV reduction initiating in Lobule X, followed by the adjacent Lobule VIIb and then expanding to Lobule VIII, Crus II, Lobules IX, Crus I, VI, IV-V and finally reaching to Lobule III. Trajectory 2 showed a reduction that started in Lobule III, gradually expanded to Lobules IV-V, VI, Crus I, Lobules X, IX, VIII, VIIb, and finally reached to Crus II. Trajectory 3 was characterized by originating in Lobule VIIb, followed by Lobule VIII, gradually expanding to Crus II, Lobule IX, Crus I, Lobule X, Lobules VI, IV-V and finally reaching to Lobule III. Patients assigned to the three subtypes did not differ significantly in sex distribution, age, education, illness duration or ancestry (all p > 0.05).

**Figure 1.**
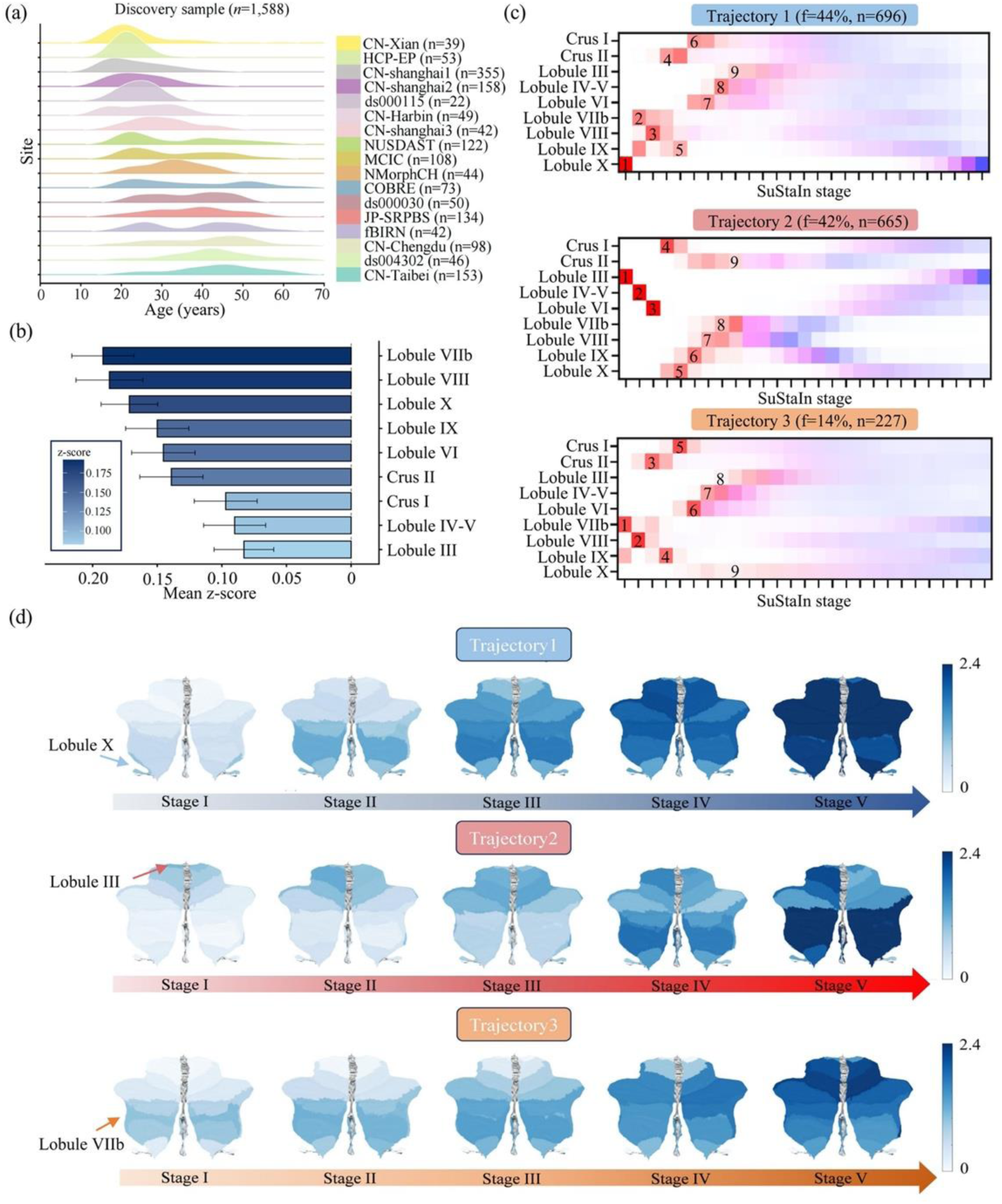
SuStaIn trajectories of cerebellar volume reduction in SCZ. **a,** Age distribution across sites in discovery sample. **b**, Mean z-scores of GMV reduction for nine cerebellar subregions. Error bar represents standard error. **c**, Sequence of spatial progression of the nine subregions estimated by SuStaIn. The positional variance diagrams visualize the cumulative probability that each subregion has reached a particular z score using different colours. Arabic numbers 1– 9 indicate the order of GMV reduction. The value of f is the proportion of individuals assigned to each trajectory. **d**, Spatiotemporal pattern of SuStaIn trajectories. Mean z scores within each SuStaIn stage are mapped to a standard cerebellar template for visualization. Color bar represents z-score. A larger z-score indicates a higher degree of GMV reduction.

### Trajectories were repeatable in two independent samples

Consistent with the trajectories from the discovery data, SuStaIn on the independent validation sample 1 (1,334 individuals with schizophrenia, 509 females, age = 36.6 ± 11.8 years; 2,652 HC, 1,423 females; age = 34.4 ± 13.3 years) and independent validation sample 2 (628 individuals with schizophrenia, 286 females, age = 24.1 ± 7.0 years; 778 HC, 397 females, age = 25.0 ± 4.3 years) replicated the three distinct and stable trajectories of cerebellar volume reduction, which originated in Lobule X, Lobule III and Lobule VIIb (**Extended Figure 2**).

### Subtype-specific Brain Morphological Patterns

In each subtype, regions of interest (ROI)-wise z-scores of GMV were mapped to a cerebellar template for visualization (**Figure 2a**). Cerebellar subregions with the greatest reduction in GMV were located at Lobule X (Left, Cohen’s d=0.61, *p*<1.1×10⁻¹⁰), Lobule III (Left, Cohen’s d=0.63, p<1.1×10⁻¹⁰), and Lobule VIIb (Left, Cohen’s d=0.83, *p*<1.1×10⁻¹⁰), respectively observed in subtype 1, 2, and 3, compared with the HC group (**Supplementary Table 3**). Cohen’s d maps from inter-subtype comparisons are visualized in **Figure 2b-c**, suggesting a severe reduction in cerebellar GMV in subtype 3 (Cohen’s d=0.80 for the inter-subtype difference at Lobule VIIb) (**Supplementary Table 4-5**). Furthermore, brain morphological analysis of the cortex and subcortex revealed that subtype 3 exhibited clear and widespread reductions in cortical thickness compared to the other two subtypes (**Extended Figure 3** and **Supplementary Data**). These results suggest that subtype 3 may represent a specific brain phenotype that is associated with pronounced neuroanatomical changes in pathological processes such as brain atrophy.

**Figure 2.**
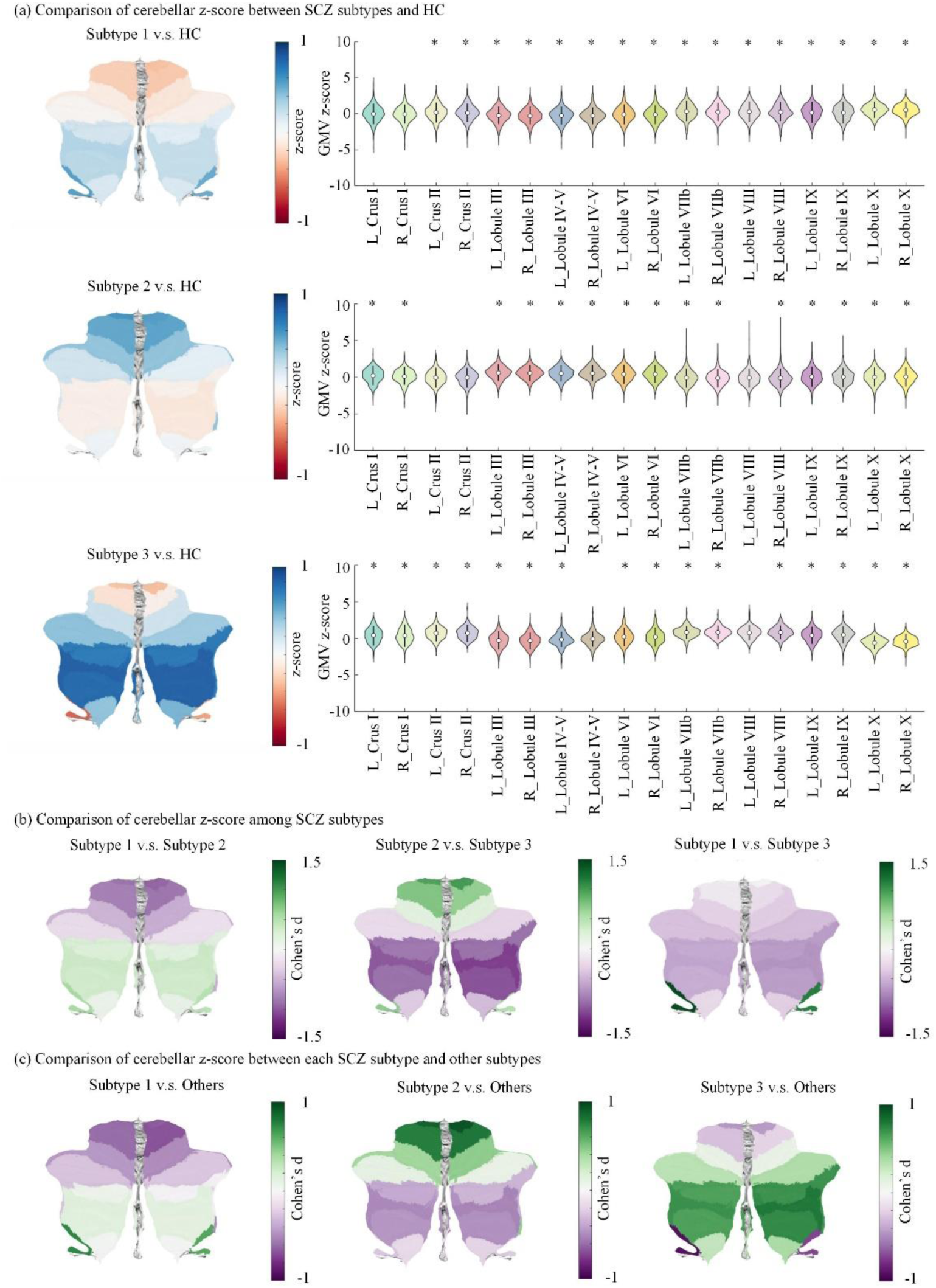
Subtype-specific pattern in cerebellar GMV reduction in SuStaIn subtypes. **a**, Reduction degree (i.e., z-score) of regional GMV is mapped to a cerebellar template for visualization. A higher z-score (colored by blue) represents a larger GMV reduction. Asterisks in the right panel indicate statistical significance (\**p*<0.001, Bonferroni correction). **b**, Comparison of cerebellar z-score among subtypes. **c,** Comparison of cerebellar z-score between each subtype and other subtypes. Analysis is performed by using two-sided two-sample *t* test. A higher Cohen’s d value (colored by green) represents a larger GMV reduction. SCZ, schizophrenia; HC, healthy controls.

### Subtype-specific Brain Functional Connectivity Characteristics

For each subtype, we characterized the whole-brain functional connectivity pattern with the origin region as the seed. The three subtypes exhibited unique and distinct connectivity patterns (**Extended Figure 4**). In brief, positive connectivity pattern in dorsal attention network (DAN) was observed only in subtype 1 and subtype 3. In addition, the positive connectivity pattern in visiual network (VIS) and negative connectivity pattern in DAN were specific to subtype 2. We also mapped the whole-brain functional connectivity pattern to a brain Automated Anatomical Labeling (AAL) atlas for visualization (**Supplementary Figure 1**).

### Subtype-specific Gene Expression Characteristics

We examined the spatial association between gene expression patterns in the brain and cortical volume alteration patterns (case-control *t* map) for each subtype. Partial least squares (PLS) analysis identified that the first PLS component (PLS1) map was spatially correlated with the *t* map for subtype 1 (*r* = 0.55, *p_spin_* = 0.014) and a trend toward significance for subtype 2 (*r* = 0.57, *p_spin_* = 0.064) (**Figure 3a**). We ranked genes contributing to PLS1 based on their weights (i.e., Z value), resulting in a total of 1125 (635 positive and 490 negative) significant genes in subtype 1, and 801 (281 positive and 520 negative) genes in subtype 2 (False Discovery Rate [FDR] correction, *p* < 0.05) (**Figure 3b** and **Supplementary Data**). Of these genes, 12 overlapped with the results from a prior schizophrenia GWAS study^[33]^ (**Figure 3c**). In addition, there were 771 genes overlapping between the two subtypes (**Figure 3d**). Gene enrichment analysis revealed that genes in subtype 1 were mainly enriched in biological processes related to metabolic signatures, which were visualized as a network diagram using Cytoscape (**Figure 3e**). In contrast, genes in subtype 2 were enriched in functional processes related to immunologic signatures (**Figure 3f**). Enrichment analysis with regard to disease ontology further revealed that the associated genes were enriched in cardiovascular and metabolic diseases in subtype 1 but in foot diseases in subtype 2 (**Figure 3g**).

**Figure 3.**
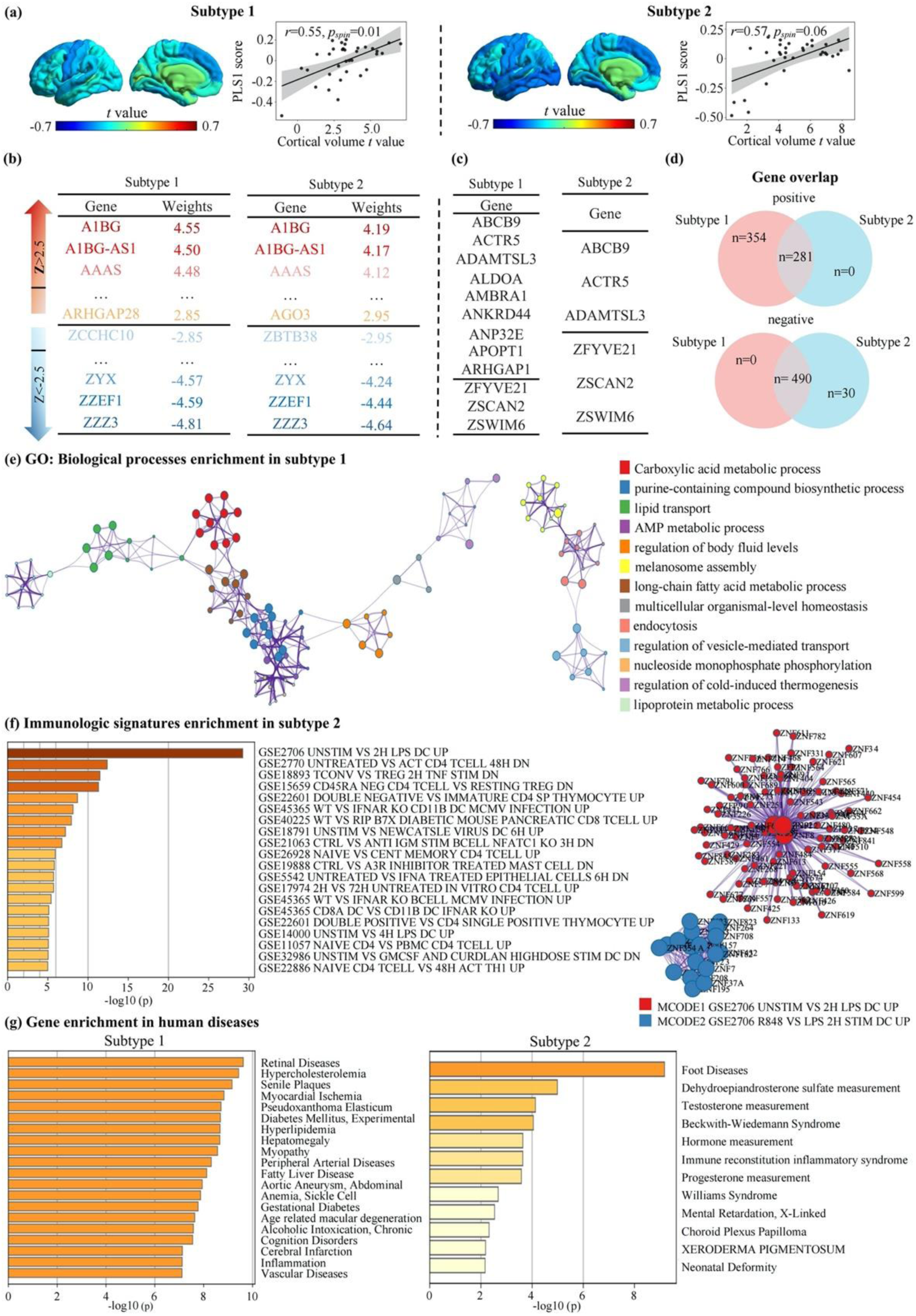
Gene expression characteristics in subtypes. **a**, Spatial association between gene expression pattern in brain and cortical volume alteration pattern (*t* map) in subtype 1 and subtype 2. Spatial pattern of the first component (PLS1) of gene expression level is mapped to a brain template for visualization. **b**, Genes with significant contribution (FDR, P < 0.05) to the PLS1 component in subtype 1 and subtype 2. **c**, Genes overlapping with SCZ GWAS result. **d**, Genes overlapping between subtype 1 and subtype 2. **e**, Top gene ontology in biological processes by enrichment analysis in subtype 1. The colors of the circles indicate different biological processes, whereas sizes reflect the number of genes. **f**, Top gene ontology in immunologic signature by enrichment analysis in subtype 2. Left panel shows the top enriched immunologic signatures, while right panel shows protein-protein interaction (PPI) network visualization. **g**, Top gene ontology in human diseases by enrichment analysis in subtype1 and subtype 2. The Benjamini–Hochberg procedure was used for multiple comparison adjustments.

### Subtype-specific Phenotypic Characteristics

There were subtype-specific associations between GMV reductions in the cerebellum and clinical symptoms (*p*<0.05) (**Supplementary Table 6**). Only in subtype 1, GMV z-scores were significantly negatively correlated with the positive and general psychopathology subscales (**Figure 4a**), indicating a lower burden of non-negative symptoms in individuals with less GMV. Only in subtype 3, we found positive correlations between less GMV and worse symptoms in cognitive and negative dimensions, both derived from the Positive and Negative Syndrome Scale (PANSS) (**Figure 4a**). In addition, analysis of variance (ANOVA) revealed significant differences in depression (*F*=3.80, *p*=0.023) and cognitive dimension (*F*=4.73, *p*=0.009) among the three subtypes (**Supplementary Table 7**). Post-hoc test indicated the most serious depression symptom in subtype 1 and cognitive symptom in subtype 3 (**Figure 4b-c**). We also found a significant correlation between SuStaIn stages and the average z-scores of cerebellar subregions in the three subtypes (subtype 1, *r*=0.98, *p*<1.0×10⁻^15^; subtype 2, *r*=0.95, *p*<1.0×10⁻^15^; subtype 3, *r*=0.95, *p*<1.0×10⁻^15^) (**Figures 4d-f**). **Supplementary Figure 2 and Supplementary Table 8** show the correlation between z-scores of each cerebellar subregion and SuStaIn stages.

**Figure 4.**
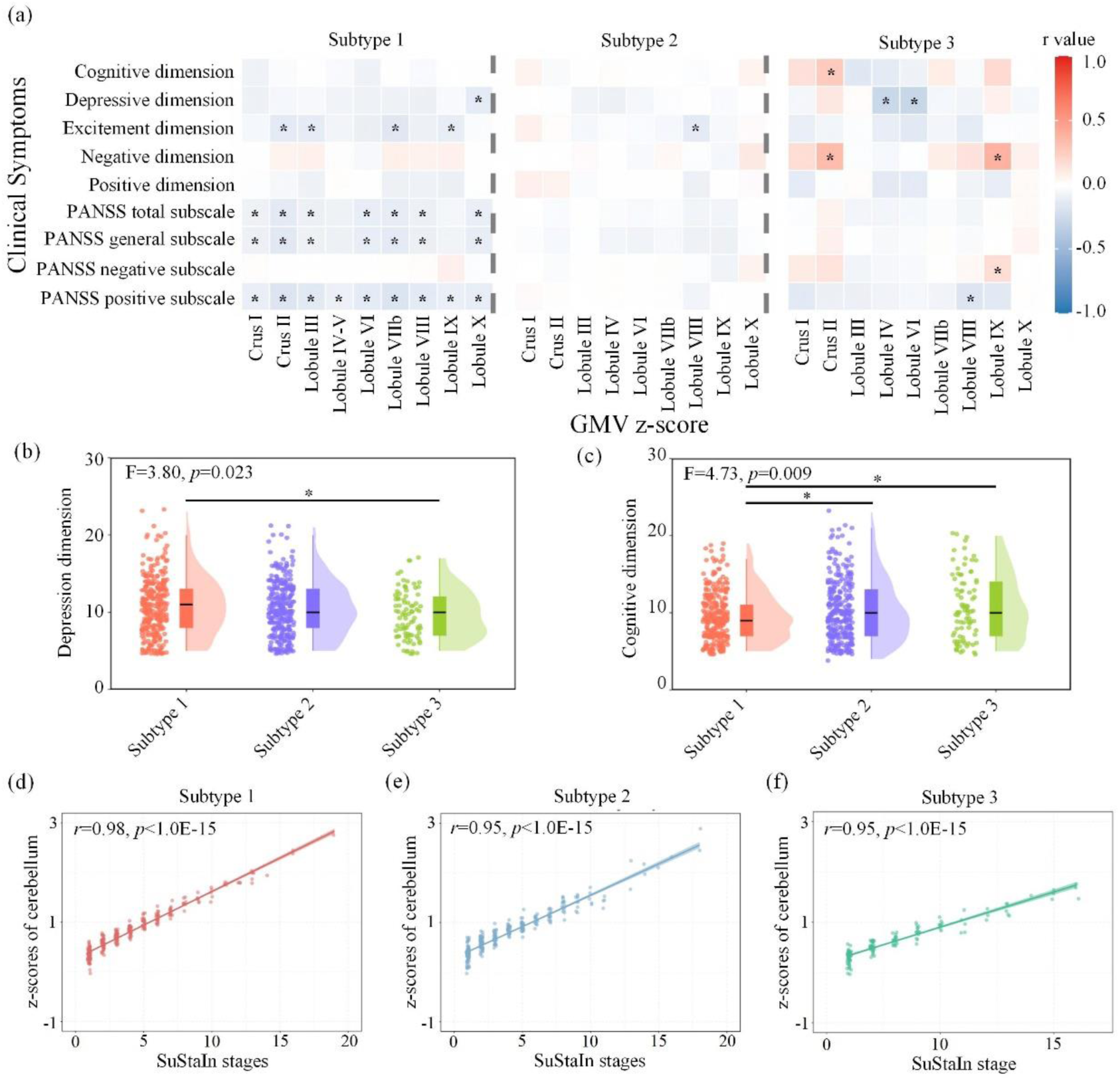
Distinct clinical profiles among subtypes. **a**, Correlation between cerebellar subregional GMV reduction (z-score) and PANSS symptoms in each subtype. Color bar represents the Spearman correlation coefficient. Asterisks indicate statistical significance (\**p*<0.05). **b-c**, Differences of symptoms in depression dimension and cognitive dimension among the three subtypes (subtype 1, N=285; subtype 2, N=294; subtype 3, N=87). One-way analysis of variance (ANOVA) and post hoc *t* test (two-tailed *p* value) are performed. Data are presented through box plots (centre line: median; bounds of box: 25th percentile to 75th percentile; whiskers: maximum and minimum value of the data). **d-f**, Correlation between the SuStaIn stages and mean z-scores of cerebellar GMV in each subtype. Spearman correlation test is performed.

### Subtype-specific association of cerebellum with APM outcomes

The APM subsamples were classified into three cerebellum subtypes according to cerebellum SuStaIn on their baseline MRI data (subtype 1, N=120; subtype 2, N=127; subtype 3, N=49) (**Supplementary Table 9**). A significant correlation between the average z-score of cerebellar subregions and follow up PANSS reduction ratio was found only in the subtype 1 (*r*=-0.21, *p*=0.026), but not in the other subtypes (subtype 2: *r*=-0.06, *p*=0.504; subtype 3: *r*=-0.08, *p*=0.600) (**Figure 5a**). Only in subtype 1, z-scores of GMV in Lobules III, IV-V and VIII were negatively correlated with follow up reduction ratio in PANSS positive, general and total scores (*p*<0.05; **Supplementary Figure 3**). Correlation analyses for each cerebellar subregion are provided in **Figure 5b** and **Supplementary Data**.

**Figure 5.**
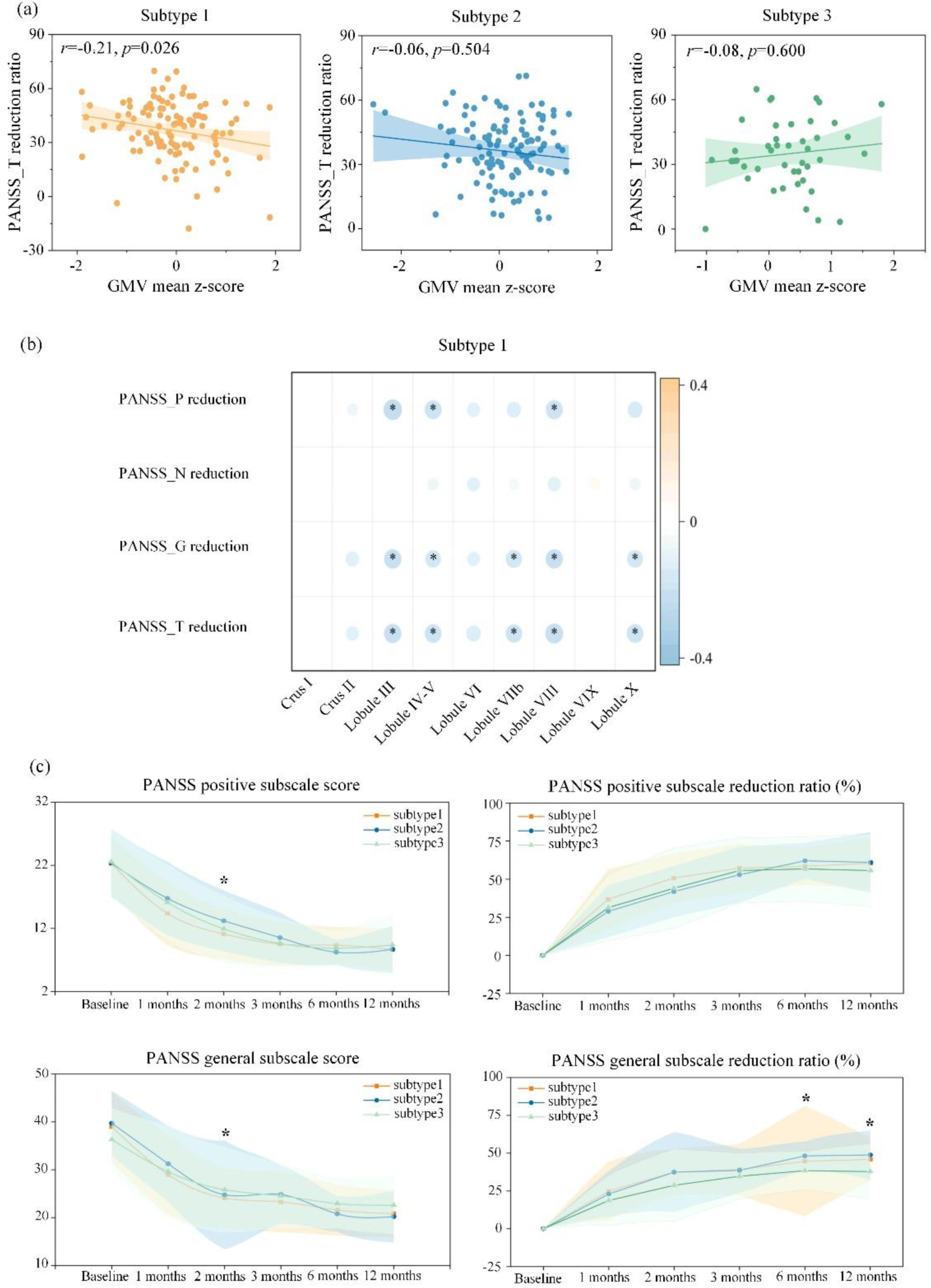
Association of subtypes with treatment outcome in APM subsamples. **a**, a significant correlation between the average z-score of cerebellar GMV and follow up PANSS reduction ratio only in the subtype 1. **b**, Correlation of each cerebellar subregion with symptom reduction ratio in PANSS **P**ositive subscale, **N**egative subscale, **G**eneral subscale, and **T**otal score. Color bar represents correlation coefficient. Asterisks (*) indicate statistical significance (*p*<0.05). **c,** Longitudinal trajectories of symptom change after the first-episode APM in the three subtypes. Asterisks (*) indicates significant difference among the three subtypes (FDR *p* < 0.05).

**Figure 6.**
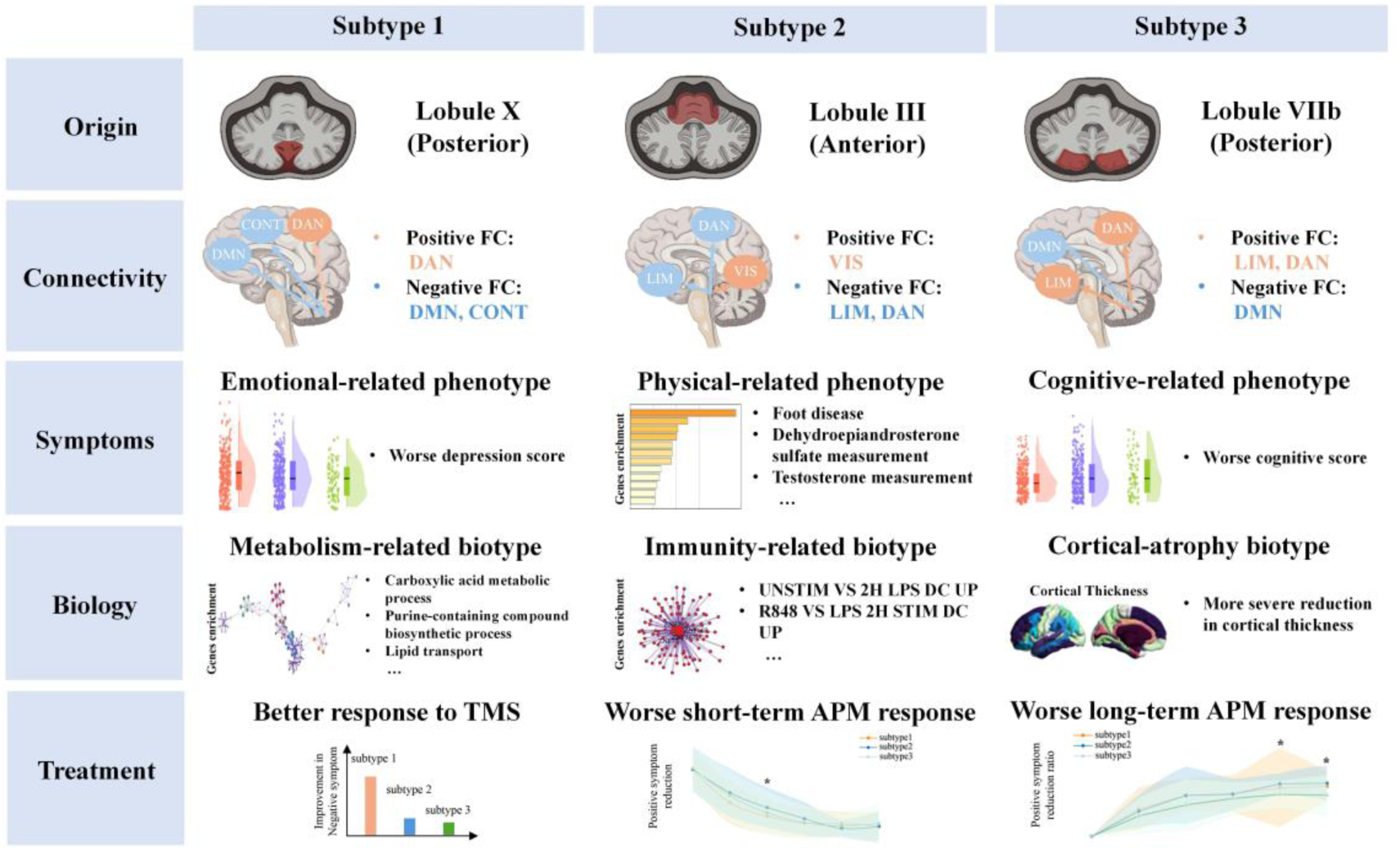
Summary of three cerebellum biotypes in SCZ.

### Subtype-specific trajectories of clinical symptoms

In the APM subsample (Beijing cohort) with a follow-up period of up to 12 months, we characterized subtype-specific trajectories of clinical symptoms after first-episode medications (**Supplementary Materials**). Longitudinal trajectories of symptoms showed that subtype 1 had a better improvement in the PANSS positive subscale at the 2 months follow up, compared to other subtypes (*F*=4.56, *p*=0.035), indicating a faster response to APM for the subtype 1 (**Figure 5c**), especially for PANSS items on delusion (*p*=0.033) and exaggeration (*p*=0.026) (**Supplementary Figure 4**). At 12-month follow up, subtype 3 showed a lesser improvement in the general subscale compared to other subtypes (*F*=4.48, *p*=0.038), suggesting a worse long-term response (**Figure 5c**), especially for PANSS items in depression (*p*=0.017) and active social avoidance (*p*=0.025) (**Supplementary Figure 4**).

### Subtype-specific association of cerebellar patterns with TMS outcomes

A subsample including 85 individuals with schizophrenia who received TMS was classified into subtype 1 (N=36), subtype 2 (N=34) and subtype 3 (N=15) (**Supplementary Table 10**). In all individuals with schizophrenia, there was no significant correlation between TMS outcomes and average z-scores of cerebellar GMV (*p*>0.05). Only in the subtype 1, we found a significant correlation between TMS outcomes (PANSS negative subscale reduction ratio) and the baseline GMV z-scores in Crus I (*r*=0.42, *p*=0.011) and Lobule IX (*r*=0.41, *p*= 0.014) (**Supplementary Figure 5**).

### Association of SuStaIn stage and treatment outcomes

We found a significant association of SuStaIn stages with PANSS symptom improvement at follow up only in the subtype 1. Specifically, the SuStaIn stages were negatively correlated with medication-treated PANSS reduction ratio in positive subscale score (*r*=-0.19, *p*=0.045) and total score (*r*=-0.21, *p*=0.029) (**Extended Figure 5a**). Furthermore, patients were classified to two substage group based on their SuStaIn stage (pre-stage I and post-stage I). We compared the differences in follow-up PANSS reduction ratio between two SuStaIn stage subgroups (pre-stage I and post-stage I). The pre-stage I group showed better APM outcomes in PANSS general subscale (Cohen’s d=0.58, *t*=2.79, *p*=0.0063) and total score (Cohen’s d=0.57, *t*=2.77, *p*=0.0065), compared to the post-stage I group (**Extended Figure 5b**).

## Discussion

Although there have been many reports on schizophrenia subtypes based on neuroimaging and clinical data, little is known about the specific pathological progression patterns in the cerebellum. Here, using the SuStaIn algorithm on cross-sectional data, we identified three distinct pathophysiological trajectories of cerebellar GMV reduction in schizophrenia, indicating that cerebellar pathology progresses along multiple subtype-specific progression patterns, which were inferred cross-sectionally. Specifically, the three trajectories map systematically onto the anterior-to-posterior functional gradient of the cerebellum: subtype 2, originating in anterior Lobule III, aligns with sensorimotor circuitry and is associated with immunological processes and neurological soft signs; whilst subtypes 1 and 3, rooted in the posterior cerebellum (Lobule X and Lobule VIIb respectively), are associated with affective and cognitive dysfunction — consistent with the established functional specialisation of posterior cerebellar regions for emotion regulation and higher-order cognition ^[19, 20, 34]^. Taken together, these findings emphasize the significance of the cerebellar subtypes of schizophrenia for clinical stratification, which also supports the biological plausibility and therapeutic relevance of the cerebellar subtypes.

Importantly, the cerebellar trajectories that we identified do not reflect a simple partition of the pathophysiological process in schizophrenia, but rather capture robust progression patterns ^[32]^. The high correlation observed in each subtype between the SuStaIn stages and the cerebellar volume reduction supports the internal consistency of the identified trajectories. This indicates that the inferred stages reflect a continuous progression of cerebellar volume reduction. Furthermore, the replication of the three trajectories in our independent validation samples also reinforces the reliability of this result. Notably, the distribution proportion of the three cerebellar subtypes remained consistent across all neuroanatomical subtypes previously defined in our work ^[35]^, with no significant interaction observed between them (*p*>0.05). This suggests that neuroanatomical and cerebellar trajectories may follow relatively independent patterns of neurostructural progression. Altogether, our findings support the view that cerebellum follows several distinct and reproducible progresses of pathological processes.

These subtype-specific patterns suggest that cerebellar pathology can be further differentiated along the existing functional axis, extending from primary sensorimotor regions to higher-order cognition systems ^[20, 36]^. In our study, subtype 1, which originates in the posterior cerebellum (Lobule X), exhibits worse affective symptoms. This is consistent with prior work reporting that the posterior cerebellum is involved in non-motor functions, particularly in affective processes ^[19, 21]^. Notably, Awada et al. recently identified a reproducible anti-coactivation pattern between the cerebellum and the ventral tegmental area, localized to bilateral paravermal Crus I/II, a posterior cerebellar region topographically close to lobule X ^[9]^. The persistent reduced frequency of this anti-coactivation pattern was associated with more severe apathy symptoms in schizophrenia, including asociality and anhedonia. Furthermore, within the cognitive dissonance framework, posterior cerebellar dysfunction is mechanistically related to reward circuit dysregulation and dopaminergic abnormalities. This provides a plausible explanation for both the more serious affective symptoms and the better early APM response observed in subtype 1. In addition to local and circuit cerebellar abnormalities, prior studies show that the connectivity between external and internal networks is also related to affective characteristics ^[37, 38]^. This is consistent with our finding that the Lobule X in subtype 1 is connected positively with DAN but negatively with DMN. By contrast, subtype 3 exhibits a more severe cognitive symptom. Existing evidence has associated the posterior cerebellum with abnormal connectivity in higher-order cognitive network ^[18]^. Given its positive connectivity with the higher-order cognitive network, subtype 3 may be related to pathological processes in cognitive function. Lastly, GMV reduction in subtype 2 originates at anterior lobe. Prior studies found that anterior lobe is connected with the primary sensorimotor cortex ^[20]^. This may suggest some of the pathological processes related to neurological soft signs (NSS), usually observed in individuals with schizophrenia ^[39, 40]^. In summary, the three subtypes further support the heterogeneity of cerebellar pathology at the functional network level.

As for biological plausibility of the three subtypes, we were able to demonstrate specific pathophysiological profiles for each subtype. Subtype 1 is characterized by the enrichment of metabolism-related genes. Although bioenergetic abnormalities are widespread in schizophrenia ^[40]^, our findings suggest that they may play a role in the pathophysiological process of this subtype ^[42, 43]^. By contrast, our findings indicate that subtype 2 shows an enrichment of immune-related genes. Prior stratification studies identified a stable immune-related subtype in schizophrenia ^[29]^, the convergence of which with our transcriptomic findings is notable. This further adds an important spatial dimension: immune-related gene expression is localised to a trajectory originating in the anterior cerebellar lobe: the phylogenetically older and ontogenetically earlier-developing region of the cerebellum. This raises the question of whether immune-mediated pathological processes, whether involving microglial activation, neuroinflammatory cascades, or aberrant developmental pruning, may disproportionately target early-developing cerebellar circuitry in a distinct subgroup of individuals with schizophrenia. The enrichment of subtype 2-associated genes in foot diseases was found via disease ontology analysis. This may have mechanistic interpretation: the anterior cerebellar lobe is the primary cerebellar substrate for sensorimotor coordination of lower-limb movement, and this enrichment likely reflects the functional anatomy of the region rather than a direct pathological association with peripheral pathology. Thus, these findings regarding the two subtypes are biologically suggestive or hypothesis-generating. Unlike the other two subtypes, subtype 3 shows the most severe brain volume reduction. Recent works have linked brain structural abnormalities to cognitive impairment ^[6, 11]^. Subtype 3 may represent a more severe pathological trajectory associated with cognition function. In conclusion, the three subtypes are associated with distinct biological processes, further supporting the new classification of biological subtypes.

We noted that treatment responses in schizophrenia may depend on subtypes and stages. Rapid symptoms improvement by APM treatment in subtype 1 is consistent with stage-dependent treatment. This finding aligns with prior evidence that during the early stages of the disease, clinical symptoms often show improvement through APM treatment, particularly in individuals with first episode schizophrenia ^[44, 45]^. Meanwhile, our results on TMS treatment have also shown evidence for efficacy in negative symptoms of subtype 1 in the later stages. Reportedly, TMS may improve negative symptoms in schizophrenia ^[46]^; concurrently, a recent study confirmed that cerebellar TMS can enhance the FC between the vermis and pallidum, suggesting functional linkage between cerebellar neuroregulation and the reward circuit ^[9, 47]^. Given that subtype 1 originates from lobule X adjacent to the vermis and is accompanied by more severe affective symptoms, these features provide a reasonable neurobiological basis for the better therapeutic effect of TMS. Together, these findings suggest that the primary therapeutic targets for schizophrenia may vary across disease progresses in subtype 1. By contrast, subtype 3 aligns with a trajectory characterized by poor longitudinal treatment outcomes in later stages. As a core and persistent feature of schizophrenia, cognitive function showed limited improvement under APM treatment ^[48]^. This phenomenon is also consistent with the neurobiological characteristics observed in treatment resistant group in prior study ^[49]^. Further longitudinal research is needed to determine the optimal timing for treatment intervention in this subtype. The clinical differences among subtypes further reveal the heterogeneity of the cerebellum in schizophrenia, which provide important support for stratified treatment strategies.

Although our work provides new insights into cerebellar pathology in schizophrenia, some limitations should be acknowledged. First, the trajectories of cerebellar GMV reduction inferred by the SuStaIn algorithm are based on cross-sectional data, which does not directly reflect the actual disease progression in patients. Future work will need to validate the reliability of this finding using more longitudinal MRI cohorts. Secondly, our spatial transcriptomics analysis relied on the AHBA, which originated from a limited number of post-mortem brain tissues from healthy adults. The AHBA analysis provides a spatial correspondence between subtype-related neuroanatomical alterations and normative gene-expression patterns, rather than direct transcriptomic evidence from patients in each subtype. Therefore, the spatial associations observed in this study still require validation using patient-level genetic data stratified by subtype. Data came from multiple different cohorts, requiring harmonization methods ^[50]^ to reduce variability across different sites, although we have ruled out the influence of site on the findings. Patients receiving TMS also received APM treatment. A further limitation of the TMS analysis is the heterogeneity of stimulation targets across the two contributing cohorts: the Xijing sample received low-frequency rTMS directed at the left temporoparietal junction (TPJ), while the Harbin sample received stimulation at the right orbitofrontal cortex (OFC). These represent mechanistically distinct targets with different postulated therapeutic mechanisms, and their pooling introduces real ambiguity regarding which neural circuit underlies the subtype 1-specific treatment benefit. Future controlled trials testing the cerebellar subtype-specific TMS hypothesis should prespecify a standardised stimulation target (ideally a posterior cerebellar or frontal site with established connectivity to reward-related circuitry) and incorporate sham controls across all three subtypes to support causal inference.

## Conclusion

This study describes three reproducible spatiotemporal trajectories of cerebellar GMV reduction in schizophrenia, each with distinct cross-sectionally inferred sequences of regional involvement. Multi-modal, transcriptomic and treatment-related analyses revealed subtype-specific biological characteristics, which provide interesting but more exploratory biological and clinical characterization. Overall, our results support biologically informed stratification in schizophrenia and may help in developing future targeted therapeutic strategies.

## Methods

### Sample Dataset

#### Cross-sectional discovery sample

The discovery sample included cross-sectional 3D T1-weighted volumetric brain MRI scans from 1,588 individuals diagnosed with schizophrenia (638 females; mean age: 31.9 ± 12.0 years) and 2,341 HC (1,041 females; mean age: 33.3 ± 13.6 years), after a standardized quality control (QC) protocol (**Supplementary Materials**). All individuals with schizophrenia were from Chinese hospitals and open datasets^[22, 35, 51–60]^. Patients were diagnosed according to the Diagnostic and Statistical Manual of Mental Disorders, fourth edition (DSM-4) or fifth edition (DSM-5). **Supplementary Tables 1 and 2** provide comprehensive demographic and clinical characteristics, along with inclusion and exclusion criteria for each cohort. Ethical approval was obtained from the Medical Research Ethics Committees of Fudan University (FE222711) and written informed consent was provided by all participants.

#### Cross-sectional validation sample

The validation sample 1 included 1,334 individuals with schizophrenia (509 females; age = 36.6 ± 13.8 years) and 2,652 HC (1,423 females; age = 34.4 ± 13.3 years) recruited from 13 cohorts of ENIGMA schizophrenia working groups from various countries around the world (**Supplementary Table 1**). The validation sample 2 included 628 individuals with schizophrenia (286 females, age = 24.1 ± 7.0 years) and 778 HC (397 females, age = 25.0 ± 4.3 years) recruited from another four independent sites, separately located at Beijing, Zhengzhou, Changsha from China (**Supplementary Table 1**).

#### Treatment sample

This comprised of APM and TMS subsamples, including 381 patients with schizophrenia. The APM subsample included longitudinal data from 296 individuals with APM-treated schizophrenia (161 females; age = 23.8 ± 6.6 years) recruited from two hospitals (**Shanghai** Mental Health Center, N = 179; Peking University People’s Hospital of **Beijing**, N=117). 274 of 296 (92.6%) were drug-naive FES patients. The Shanghai cohort had a total of 179 individuals with schizophrenia (96 females; age = 25.6 ± 7.3 years) who were followed up within approximately three months (short-term subgroup). At baseline, 157 patients were treatment-naïve FES. Of these, 126 received monotherapy and the remaining 53 patients received combined therapy (≥two antipsychotic drugs). The daily dosage of drugs converted to chlorpromazine (CPZ) was 431.4 ± 217.4 mg d^-1^. At follow-up, 133 of 179 (74.3%) were considered responders whose symptom relief (percentage of reduction ratio in PANSS) >25%.

The Beijing cohort had 117 individuals with schizophrenia (65 females; age = 21.2 ± 4.0 years) who were treatment-naïve FES at baseline and followed up more than one times (up to 5 times). Of them, 80 patients were successfully followed up after 12 months (long-term subgroup). The mean CPZ during medication was 256.4 ± 143.2 mg d^-1^.

The TMS subsample included a total of 85 patients with schizophrenia, from two hospitals (Xijing Hospital of **Xi’an**, N=36; First Specialized Hospital of **Harbin**, N=49), who were treated with TMS under stable dosage of antipsychotics and included in the longitudinal analyses. At baseline, 62 of them were treatment-naïve FES. Briefly, the stimulation target was set at the left TPJ for 36 individuals and at the right OFC for 49 individuals. PANSS assessments were performed at baseline and at follow-up by the same psychiatrist. The average duration of PANSS follow-up was 3.0 weeks. At follow-up, 61 of 85 (71.8%) individuals with schizophrenia were considered as TMS responders, defined as a reduction in PANSS total score of greater than 25%.

Details of APM and TMS subjects are described in **Supplementary Materials** and **Supplementary Table 9-10**. We use the naturalistic data from TMS samples collected during routine clinical care; this is not a report of a randomized trial. The TMS study was registered in the Chinese Clinical Trials Registry (ChiCTR2000041106). The study has been approved by the Medical Research Ethics Committees of the local hospitals (ethics numbers: 2017-36R (Shanghai), 2008-2 (Beijing), XJYYLL-2015047 (Xi’an) and IRB2019-004 (Harbin)).

#### Data Processing

Brain MRI data were acquired from each study center (**Supplementary Table 2**). Structural MRI data were processed by using a standardized protocol from the ENIGMA Computational Anatomy Toolbox (CAT12; https://neuro-jena.github.io/enigma-cat12/) ^[61]^. This protocol can yield morphometric measures (i.e., GMV) at the regional level, estimated from structural images. Based on Automated Anatomical Labeling (AAL3) atlas, we extracted a total of nine ROIs from cerebellum, including bilateral Crus I, Crus II, Lobules III, IV-V, VI, VIIb, VIII, IX, and X. The ROI-wise GMV measurements of cerebellum subregions were first adjusted by regressing out the effects of sex, age, age squared, scan site and total intracranial volume (TIV), via a multiple linear regression model. These adjusted values were further converted to z-score and multiplied by-1 so that a higher z-score corresponds to a greater GMV reduction (i.e., normative deviations from the average level of HC). In addition, structural data were processed by using FreeSurfer (version 7.3; http://surfer.nmr.mgh.harvard.edu/), yielding cortical thickness, surface area, and cortical volume estimation for 68 cortical and 14 subcortical regions.

#### SuStaIn

To estimate the potential cross-sectional “trajectories” of GMV reduction within cerebellar subregions in schizophrenia, we employed a machine learning modeling (i.e., Subtype and Stage Inference [SuStaIn] ^[32]^) on ROI-wise z-score from nine cerebellar subregions. SuStaIn parameters are provided in **Supplementary Materials** and described briefly here. Input is an M×N matrix, where M corresponds to the number of schizophrenia cases (M=1,588) and N represents the number of cerebellar subregions (N=9). SuStaIn can estimate the most probable sequence of spatial progression (i.e., cross-sectional inferred “trajectory”), and further identify the potential distinct trajectories with a given subtype number k. The choice of optimal k is provided in **Supplementary Materials**. In addition, SuStaIn can achieve individualized subtyping and sub-staging by a two-step assignment. For each patient, it calculated a probability (ranging from 0 to 1) of belonging to each “trajectory” and further assigned the patient to a SuStaIn stage (ranging from 0 to 1) of the maximum-likelihood “trajectory”, through Markov Chain Monte Carlo (MCMC) iterations.

#### Subtype-specific SuStaIn Trajectory

To characterize the spatiotemporal “trajectories” of cerebellar volume reduction, we calculated the mean z-score of GMV across individuals within the same sub-stage of each SuStaIn trajectory. ROI-wise z-scores were projected onto a cerebellar surface mesh template using the Spatially Unbiased Infratentorial Template (SUIT) toolbox (https://www.diedrichsenlab.org/imaging/suit.htm). To examine whether SuStaIn stages (i.e., a continuous index of the “temporal” progression of SuStaIn “trajectory”) correlate with the pathological processes and clinical symptoms of schizophrenia, we tested Spearman correlation between the SuStaIn stages and cerebellar regional z-scores for each subtype. We also investigated the associations between SuStaIn stages and variables including age, disease duration and PANSS subscales.

### Subtype-specific Biological Characteristics

#### Morphological pattern

To visualize the cerebellar GMV reduction patterns of each subtype, the mean z-score of each cerebellar subregion was calculated to quantify the degree of deviation from HC level. To comprehensively characterize the brain morphological signatures of each subtype, morphological measures (cortical thickness, area, volume and subcortical volume) were also included. Two sample *t* test was performed to compare difference between any two subtypes. For multiple comparisons, Bonferroni correction was used for the mean z-score and FDR correction was used for the brain morphological measures.

#### Connectivity pattern

We characterized the seed-to-ROIs FC pattern in each subtype using resting-state fMRI from a subsample including 574 individuals with schizophrenia (**Supplementary Materials**). Specifically, two brain atlases (anatomical AAL and functional Schaefer) were used to define ROIs. Cerebellar Lobule X, III and VIIb were defined as the seeds because they were the first regions observed in SuStaIn trajectories. In each subtype, one sample *t* test was used to characterize the significant connectivity pattern of the seed region with other ROIs. FDR were corrected for multiple comparisons.

#### Subtype-specific Gene Expression Pattern

Brain-wide gene expression profiles and microarray expression data were acquired from the Allen Human Brain Atlas ^[62]^ (AHBA; http://human.brain--map.org). PLS regression was used to investigate the spatial association between the t map of case-control difference in the cortical volume and the gene expression profiles (**Supplementary Materials**). PLS analysis ^[63]^ was used to investigate the association between the expression levels of all 12,668 genes and case-control difference *t* map in the cortical volume for each subtype. Significant genes (FDR *p* < 0.05) were extracted for enrichment analysis using Metascape ^[64]^(v3.5).

#### Phenotypic Characteristics across Subtypes

Demographic, clinical and global brain variables available for the discovery cohort included age (*n*=1,588), disease duration (*n*=846), PANSS subscale-level (*n*=1,078) and PANSS item-level (*n*=666) (**Table 1**). To identify subtype-specific phenotypic features, statistical comparisons of these variables between the three subtypes and HC were conducted by two steps: (1) comparisons with all other subtypes (i.e., one-to-all comparisons). Each subtype was compared with all individuals in the other subtypes using a one-to-all approach to identify subgroup-specific characteristics; and (2) direct comparisons with another subtype (i.e., one-to-one comparisons). All comparisons were performed using ANOVA, with appropriate post hoc tests. We also examined the association between regional volume reduction and PANSS symptoms in each subtype.

**Table 1.**
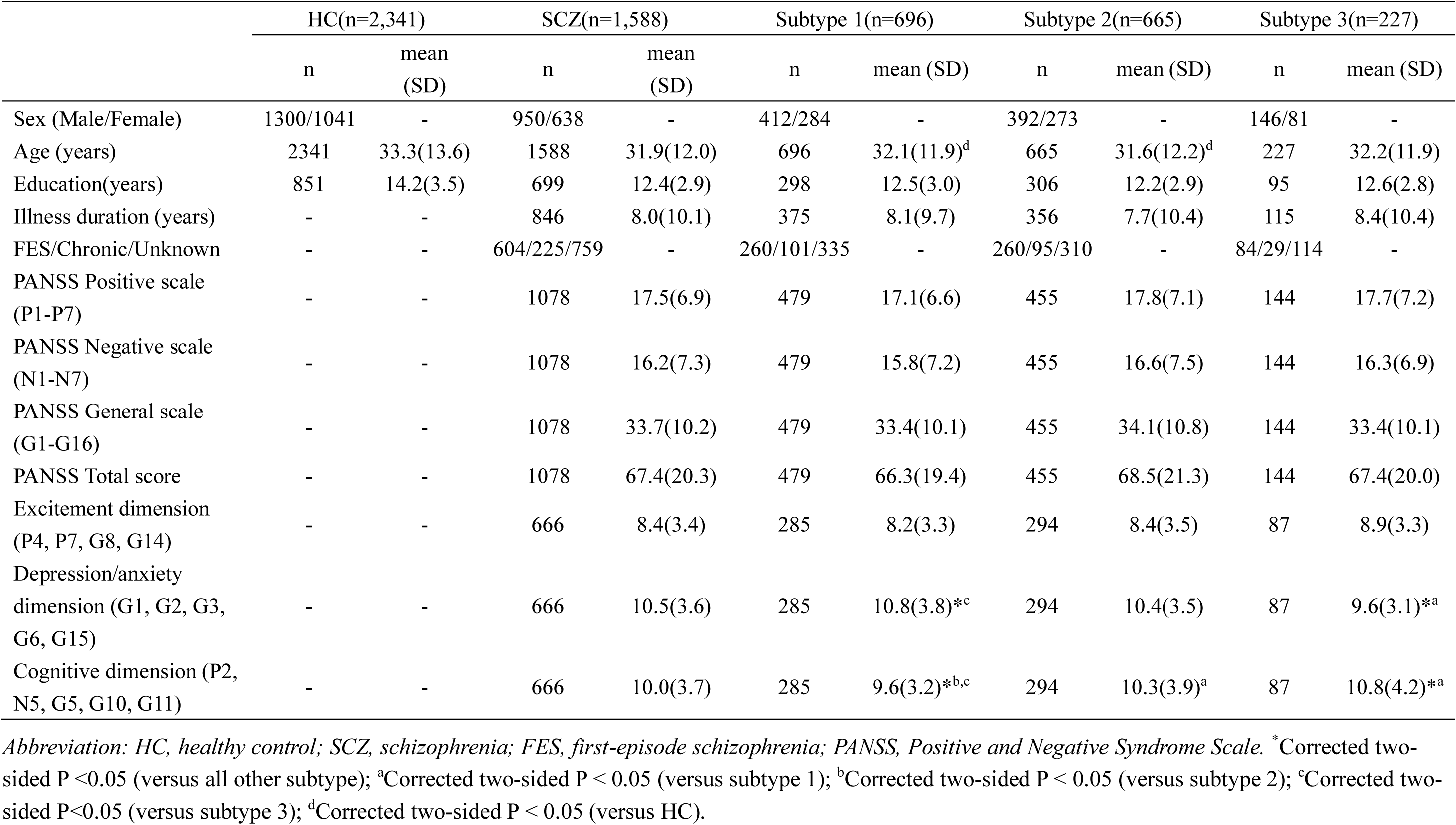
Demographic and clinical characteristics between individuals with schizophrenia and HC in discovery dataset.

|  | HC(n=2,341) |  | SCZ(n=1,588) |  | Subtype 1(n=696) |  | Subtype 2(n=665) |  | Subtype 3(n=227) |  |
| --- | --- | --- | --- | --- | --- | --- | --- | --- | --- | --- |
|  | n | mean (SD) | n | mean (SD) | n | mean (SD) | n | mean (SD) | n | mean (SD) |
| Sex (Male/Female) | 1300/1041 | - | 950/638 | - | 412/284 | - | 392/273 | - | 146/81 | - |
| Age (years) | 2341 | 33.3(13.6) | 1588 | 31.9(12.0) | 696 | 32.1(11.9)<sup>d</sup> | 665 | 31.6(12.2)<sup>d</sup> | 227 | 32.2(11.9) |
| Education(years) | 851 | 14.2(3.5) | 699 | 12.4(2.9) | 298 | 12.5(3.0) | 306 | 12.2(2.9) | 95 | 12.6(2.8) |
| Illness duration (years) | - | - | 846 | 8.0(10.1) | 375 | 8.1(9.7) | 356 | 7.7(10.4) | 115 | 8.4(10.4) |
| FES/Chronic/Unknown | - | - | 604/225/759 | - | 260/101/335 | - | 260/95/310 | - | 84/29/114 | - |
| PANSS Positive scale (P1-P7) | - | - | 1078 | 17.5(6.9) | 479 | 17.1(6.6) | 455 | 17.8(7.1) | 144 | 17.7(7.2) |
| PANSS Negative scale (N1-N7) | - | - | 1078 | 16.2(7.3) | 479 | 15.8(7.2) | 455 | 16.6(7.5) | 144 | 16.3(6.9) |
| PANSS General scale (G1-G16) | - | - | 1078 | 33.7(10.2) | 479 | 33.4(10.1) | 455 | 34.1(10.8) | 144 | 33.4(10.1) |
| PANSS Total score | - | - | 1078 | 67.4(20.3) | 479 | 66.3(19.4) | 455 | 68.5(21.3) | 144 | 67.4(20.0) |
| Excitement dimension (P4, P7, G8, G14) | - | - | 666 | 8.4(3.4) | 285 | 8.2(3.3) | 294 | 8.4(3.5) | 87 | 8.9(3.3) |
| Depression/anxiety dimension (G1, G2, G3, G6, G15) | - | - | 666 | 10.5(3.6) | 285 | 10.8(3.8)<sup>*c</sup> | 294 | 10.4(3.5) | 87 | 9.6(3.1)<sup>*a</sup> |
| Cognitive dimension (P2, N5, G5, G10, G11) | - | - | 666 | 10.0(3.7) | 285 | 9.6(3.2)<sup>*b,c</sup> | 294 | 10.3(3.9)<sup>a</sup> | 87 | 10.8(4.2)<sup>*a</sup> |

*Abbreviation: HC, healthy control; SCZ, schizophrenia; FES, first-episode schizophrenia; PANSS, Positive and Negative Syndrome Scale.* <sup>\*</sup>Corrected two-sided P < 0.05 (versus all other subtype); <sup>a</sup>Corrected two-sided P < 0.05 (versus subtype 1); <sup>b</sup>Corrected two-sided P < 0.05 (versus subtype 2); <sup>c</sup>Corrected two-sided P < 0.05 (versus subtype 3); <sup>d</sup>Corrected two-sided P < 0.05 (versus HC).

#### Association of Clinical Treatments with Subtypes

This exploratory analysis investigated whether subtype-specific information, based on only their baseline MRI data, was associated with their treatment responses at follow up. The SuStaIn model, which was trained using the discovery sample, assigned each individual with schizophrenia to one of three subtypes and further to a sub-stage based on their pre-treatment MRI data. For each individual, the SuStaIn model provided three sets of quantitative metrics in assessing cerebellar spatiotemporal progression: subtype probability score, sub-stage score and subregional z-score.

Following baseline MRI subtyping, individuals with schizophrenia received either pharmacological treatment or TMS. The treatment outcomes at follow-up were evaluated by the reduction rate of PANSS subscale score (i.e., reduction rate = [ (baseline-follow-up) / baseline] × 100%). A total of 296 pharmacologically treated individuals with schizophrenia (179 short-term follow-up sample and 117 long-term [follow-up sample) and 85 TMS (combined with medications) individuals with schizophrenia were included in the longitudinal analysis (**Supplementary Table 9-10**). Spearman correlation analysis was performed to examine correlation between the cerebellar spatiotemporal features and treatment outcomes. We compared the differences of treatment outcomes across the three subtypes using one-to-one and one-to-all statistical approaches.

In addition to the three subtypes mentioned above, we further divided individuals within each phenotype into two subgroups (pre-stage I and post-stage I) based on differences of SuStaIn stages (i.e., ‘temporal’ phenotype). Two-sample *t* test was used to compare the differences in follow-up treatment outcomes between the two subgroups in the APM and TMS samples.

## Data availability

The HCP-EP dataset is available at https://www.humanconnectome.org/study/human-connectome-project-for-early-psychosis/. The NMorphCH dataset and NUSDAST dataset are downloaded through the SchizConnect (https://www.nitrc.org/projects/schizconnect/). The COBRE dataset is download from the Center for Biomedical Research Excellence in Brain Function and Mental Illness (COBRE) (https://coins.trendscenter.org/). The FBIRN dataset is download from https://www.nitrc.org/projects/fbirn/.The DS000115 dataset is downloaded from OpenfMRI database (https://www.openfmri.org/). The DS000030 dataset is available at https://legacy.openfmri.org/dataset/ds000030/. The DS004302 dataset is available at https://openneuro.org/datasets/ds004302/versions/1.0.1. The Japanese SRPBS Multi-disorder MRI Dataset is available at https://bicr-resource.atr.jp/srpbsopen/. Requests for ENIGMA data can be applied via the ENIGMA Schizophrenia Workingdata Group (https://enigma.ini.usc.edu/ongoing/enigma-schizophreniaworking-group/). Data from other hospital-based cohorts (partial discovery dataset, cross-sectional dataset, longitudinal APM and TMS data) are not publicly available due to participant privacy and institutional data-sharing restrictions.

## Code availability

The visualization of ROI-wise z-score images was conducted using SUIT toolbox (https://www.diedrichsenlab.org/imaging/suit.htm). The visualization of functional connectivity results was achieved using the R package “ggseg” (https://github.com/ggsegverse/ggseg) and “circlize” (https://github.com/jokergoo/circlize). SuStaIn algorithm is available on the UCL-POND GitHub (https://github.com/ucl-pond/). The software and code used in T1 and functional image processing are publicly available and are described in the Methods. Statistical analyses, including correlation analysis, were conducted using MATLAB v9.7. Code for the cerebellar spatiotemporal subtyping pipeline is implemented in an open-source Python toolbox, the Cerebellar Spatiotemporal Subtyping Toolbox (CST), which is publicly available (https://github.com/Zhaoyun-Liu/CST).

## Author contributions

ZL and YJ led the project. PMT, JAT, TGME provided crucial contribution to the organization and cooperation of the project via ENIGMA. ZL and YJ take responsibility for the integrity of the data and the accuracy of the data analysis in the study. Study concept and design 1: ZL, YJ. Acquisition, analysis, interpretation of data 2: ZL, YJ, ZH, XJ, JZ, Hhu, QG, TZ, JW, WY, YZ, XY, HH, CL, RH, JM, KN, TK, FT, PU, LT, BS, IN, FS, NA, AJ, HJ, NJ, JL, UD, DG, SM, KF, RL, MZ, EP, RS, AK, GS, FP, DV, NB, ASG, ACH, KS, GHC, YQ, MJQ, YC, WK, SO, FL, GP, ADB, Sco, ANB, ATB, MC, MT, TR, SLR, MH, WW, Sca, PJS, ER, GC, SS. Drafting of the original manuscript 3: ZL, YJ. Crucial advice to the revision of manuscript and the study 4: YJ, LP, PMT, XC, YT, LC, DY, JF. All authors contributed edits and approved the contents of the manuscript.

## Data Availability

All data produced in the present study are available upon reasonable request to the authors.

https://www.humanconnectome.org/study/human-connectome-project-for-early-psychosis/

https://www.openfmri.org/

https://legacy.openfmri.org/dataset/ds000030/

---

**Extended Figure 1.**
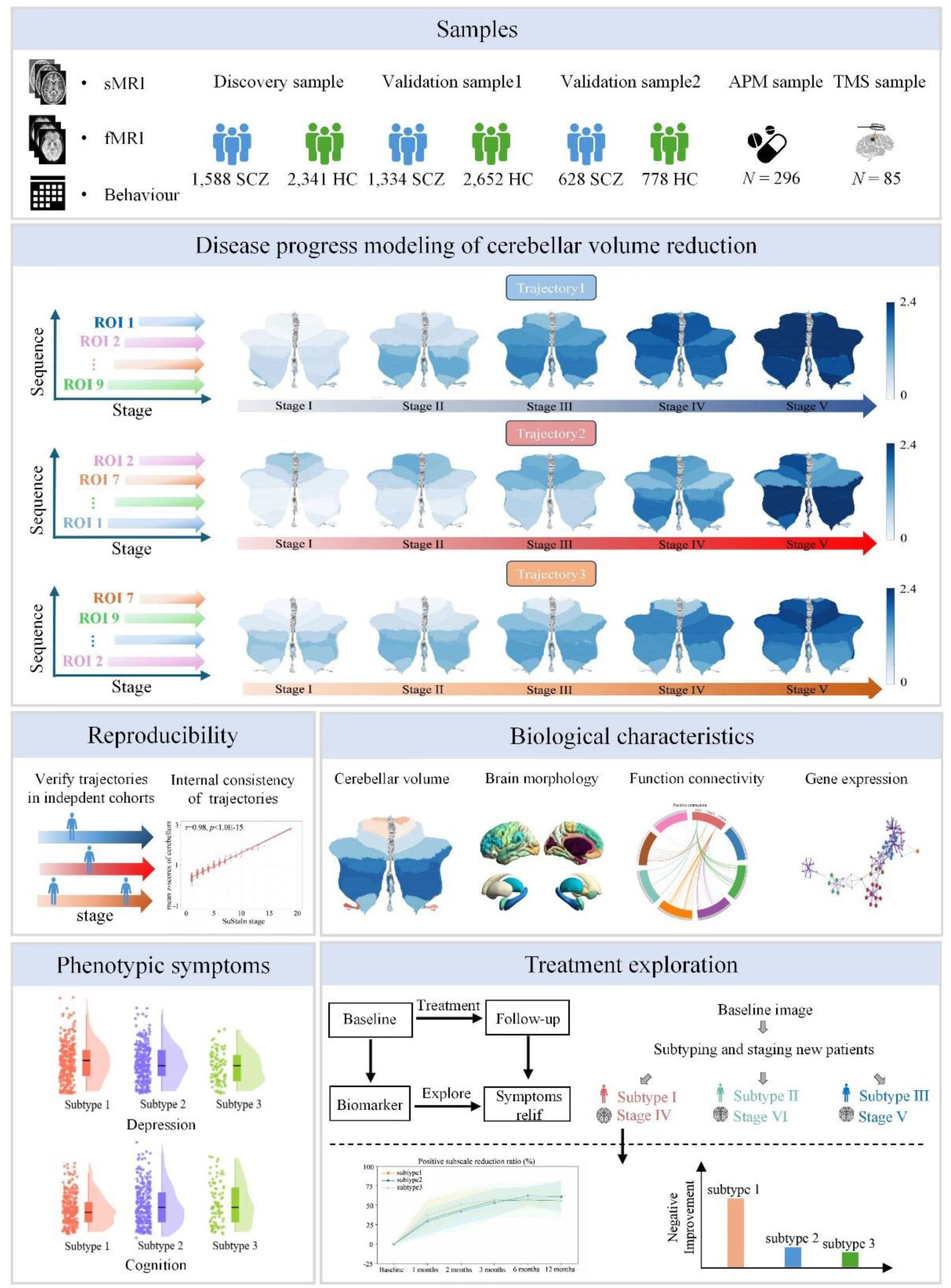
Study design and analytical framework for identifying cerebellar subtypes in schizophrenia. (Top panel) Data processing and feature extraction. Large-scale multimodal MRI data underwent standardized preprocessing to extract cerebellar gray matter volume, cortical and subcortical morphology and functional connectivity features. The first two metrics were harmonized and transformed into z-score relative to HC. Finally, all metrics construct a unified multimodal feature matrix. (**A**) Multimodal analysis. The subtype and Stage Inference (SuStaIn) algorithm was employed to disentangle the pathophysiological trajectories of cerebellar volume reduction, identifying three distinct subtypes and their progression trajectories. Subsequent analyses characterized subtype-specific alterations in cortical and subcortical morphology (thickness, area, volume) and evaluated whole-brain functional dysconnectivity using the AAL and Schaefer 2018 (400-parcel) atlases. (**B**) Clinical follow-up and treatment analysis.To validate clinical relevance, a longitudinal cohort (N=381) was mapped onto the established SuStaIn model for individualized subtyping and staging. The analysis investigated distinct clinical symptom profiles (PANSS subscales) and assessed long-term treatment outcomes in response to antipsychotic medication and repetitive transcranial magnetic stimulation. (**C**) Gene enrichment analysis. Transcriptomic signatures underlying subtype-specific volume reduction were explored using the Allen Human Brain Atlas (AHBA). Partial least squares regression was utilized to link spatial volume t maps with gene expression profiles, followed by functional enrichment analysis to identify associated biological pathways and risk genes.

**Extended Figure 2.**
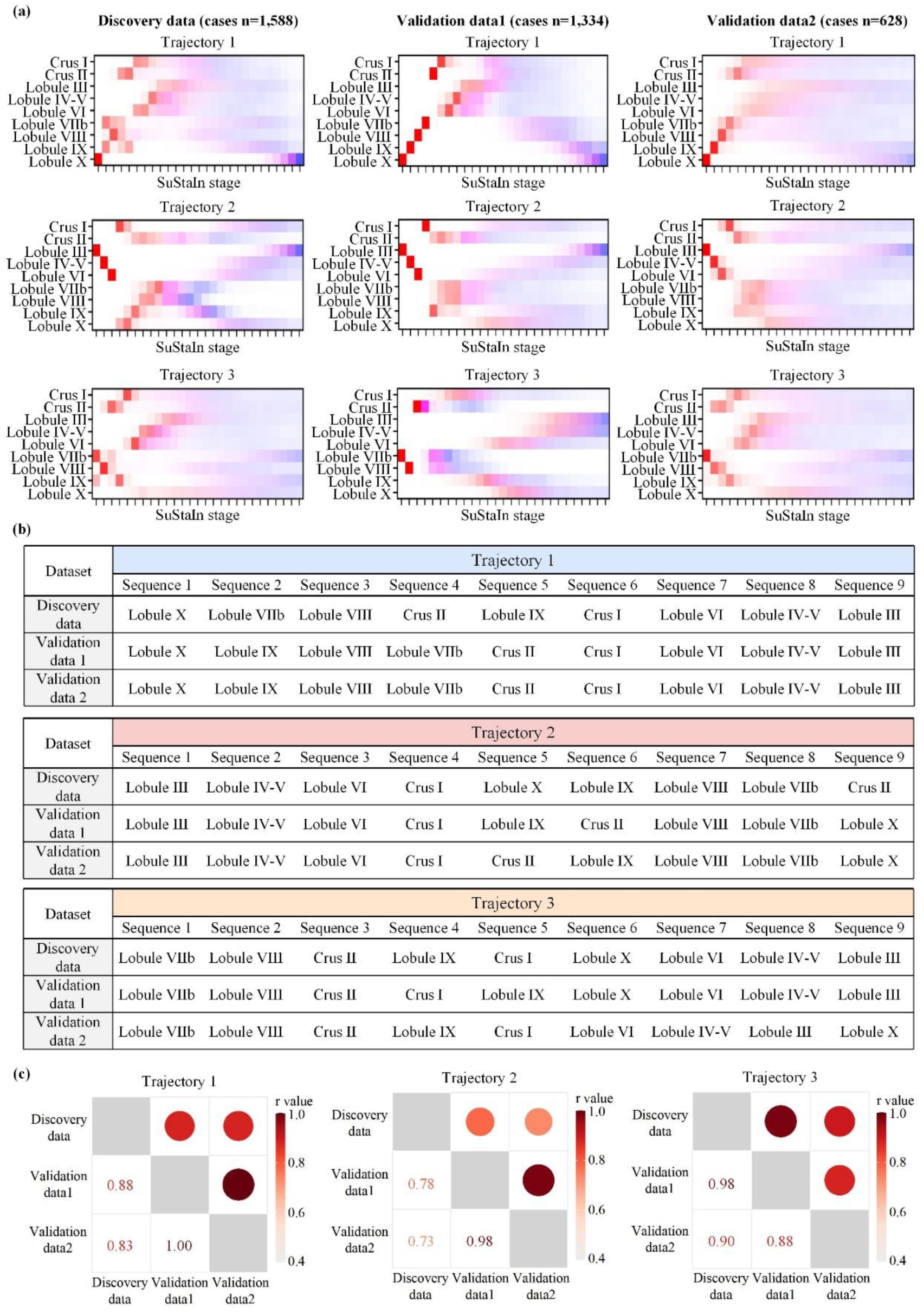
Reproducibility of cerebellar trajectories across discovery and two independent validation datasets. (**a**) SuStaIn on two independent validation samples repeats three distinct and stable trajectories of cerebellar volume reduction, which originated at Lobule X, Lobule III and Lobule VIIb. (**b**) Detailed regional ordering along each of the three trajectories in the discovery dataset and two independent validation datasets. (**c**) Cross-dataset order similarity of each trajectory using Spearman’s correlation. Separate correlation matrices are shown for Trajectories 1–3. In each matrix, circles in the upper triangle represent the magnitude of the Spearman correlation coefficient, with larger circles and darker red colors indicating greater similarity. Exact Spearman r values are shown in the lower triangle, while diagonal cells are shaded in gray to denote self-correlations.

**Extended Figure 3.**
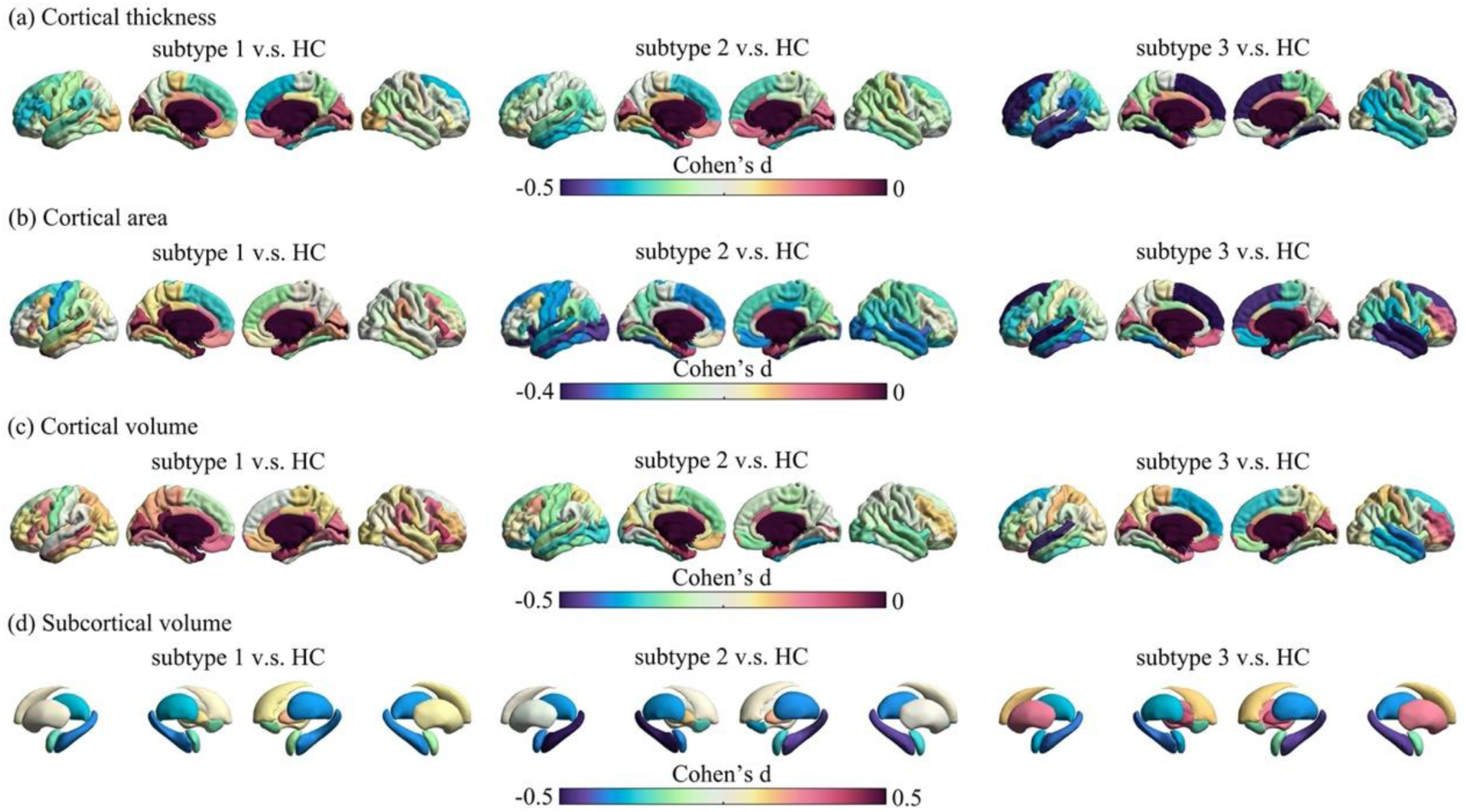
Group difference of brain morphological signatures in cortex and subcortex. Morphometric measures include (**a**) cortical thickness, (**b**) cortical area, (**c**) cortical volume, and (**d**) subcortical volume. Color bar indicates Cohen’s d value. A smaller negative Cohen’s d value indicates larger reduction of morphometric measures in cases compared with healthy control (HC) group.

**Extended Figure 4.**
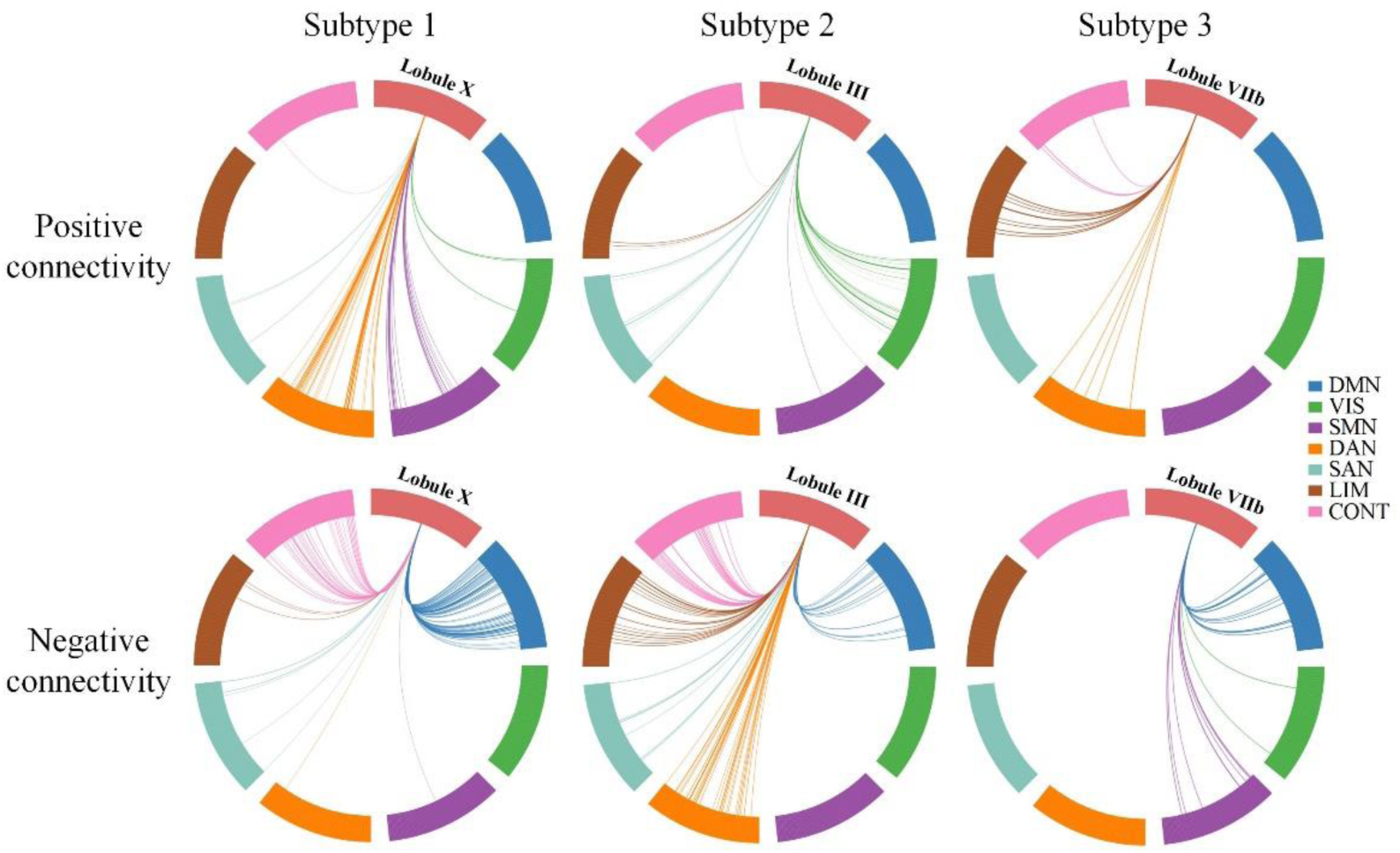
Subtype-specific functional connectivity pattern. The Circos connection plots show connectivity patterns (both positive and negative) in each of the three subtypes (corresponding to three origins: Lobule X, Lobule III, Lobule VIIb). Only connectivity with significant threshold (|*t*|>5) are displayed. Multiple comparison correction for statistical comparisons was performed using FDR correction. Whole brain consists of 400 cortical regions, which are further classified to 7 resting-state functional networks, including Visual (VIS), Somatomotor (SMN), Dorsal Attention (DAN), Salience (SAN), Limbic (LIM), Control (CONT), Default (DMN).

**Extended Figure 5.**
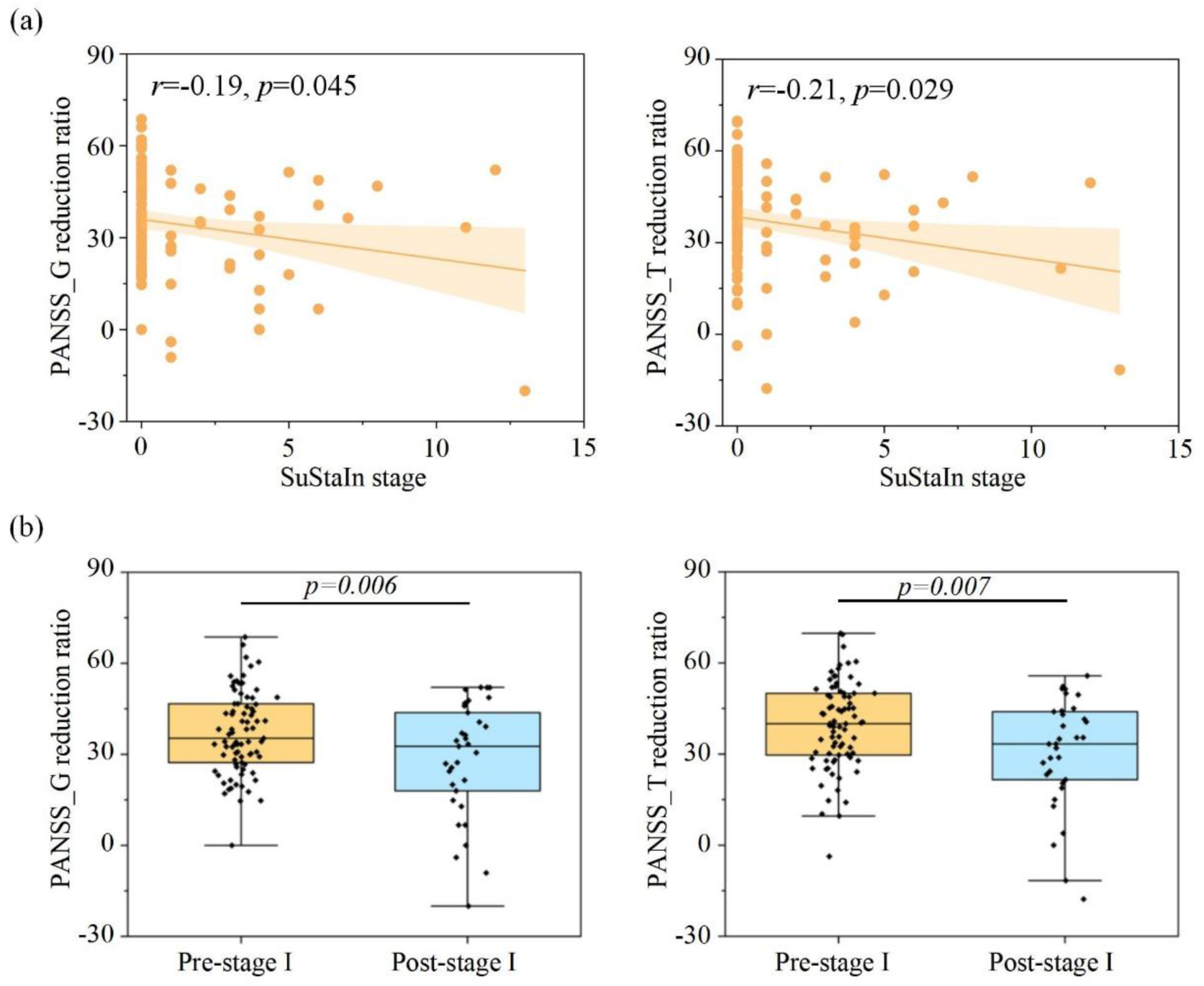
Association of SuStaIn stage and treatment outcomes in subtype 1. **a**, a significant correlation between SuStaIn stage and follow-up PANSS reduction ratio only in the subtype 1. **b**, Differences of PANSS reduction ratio among the two substage group based on their SuStaIn stage (pre-stage I and post-stage I). Two sample *t* test is performed.

